# SARS-COV-2 THREE FORCING SEASONALITIES: POLICIES, ENVIRONMENT AND URBAN SPACES

**DOI:** 10.1101/2020.07.15.20154823

**Authors:** Charles Roberto Telles

**Affiliations:** Multisector Projects Division. Secretary of State for Education and Sport of Paraná. Água Verde Avenue, 2140. Água Verde. Curitiba - PR, 80240-900. Brazil

**Keywords:** COVID-19, policies and ALE preventive methods, forced seasonality, Fourier transforms, environmental driven factors, urban spaces heterogeneity

## Abstract

This research investigated if pandemic of SARS-COV-2 follows the Earth seasonality *ε* comparing countries cumulative daily new infections incidence over Earth periodic time of interest for north and south hemisphere. It was found that no seasonality in this form *ε* occurs as far as a seasonality forcing behavior *ε*′ assumes most of the influence in SARS-COV-2 spreading patterns. Putting in order *ε*′ of influence, there were identified three main forms of SARS-COV-2 of transmission behavior: during epidemics growth, policies are the main stronger seasonality forcing behavior of the epidemics followed by secondary and weaker environmental and urban spaces driving patterns of transmission. At outbreaks and control phase, environmental and urban spaces are the main seasonality forcing behavior due to policies/ALE limitations to address heterogeneity and confounding scenario of infection. Finally regarding S and R compartments of SIR model equations, control phases are the most reliable phase to predictive analysis.

These seasonality forcing behaviors cause environmental driven seasonality researches to face hidden or false observations due to policy/ALE interventions for each country and urban spaces characteristics. And also, it causes policies/ALE limitations to address urban spaces and environmental seasonality instabilities, thus generating posterior waves or uncontrolled patterns of transmission (fluctuations).

All this components affect the SARS-COV-2 spreading patterns simultaneously being not possible to observe environmental seasonality not associated intrinsically with policies/ALE and urban spaces, therefore conferring to these three forms of transmission spreading patterns, specific regions of analysis for time series data extraction.

## 1) Introduction

The main focus of this research is to point, as noted in Grassly and Fraser [1], the consequences of seasonality for endemic *R*_0_ stability in order to understand and obtain an endemic equilibrium for COVID-19 involving mixing patterns such as environmental driving factors, policies interventions and urban spaces [3-8]. These three variables might pose a challenging outcome for predictive analysis [9] of SARS-COV-2 spreading patterns since the time series data of cumulative daily new cases are highly influenced by it in terms of quantitative outcomes day by day, fluctuations and mainly random outcomes that comes influenced by different aspects of local epidemics behavior. In order to correct and address this later point, we might be observing data that should be divided in three phases of epidemics that is the outbreak, peak and control followed by its main seasonality drivers found in this research in sequence that is the environmental variables (Earth seasons and atmospheric conditions), policies and ALE interventions and urban spaces (local indoor and outdoor spaces for transit and social interactions being public or private with natural physical features on it). By observing time series data of cumulative daily new cases worldwide [10], these three sequences of epidemics phases present different results for each sample (country) of observation and many delays in order to obtain a normality for epidemic curve are found as well as attractive behavior of the outcomes over time. These constraints give the formation of false observations of phenomenon to predictive analysis based on SIR models and derivations [11], policies interventions and the main role of environmental variables towards outbreaks and waves restarting periods.

Following this late paragraph statements, this research divided the world data of cumulative daily new cases for COVID-19 in three regions of data extraction in the linear time series, designed to organize the confounding data of analysis, prediction and accuracy for fields of research. This will bring more robust understanding for the scientific convergence of results and worldwide strategies to reduce SARS-COV-2 spreading patterns of infection.

## 2) Methodology

### 2.1 Earth seasons: undefined time intervals of analysis for periodic oscillations

To put COVID-19 under the Earth seasons aspect of analysis the endemic free-equilibrium need to be under the view of Floquet Theory as it is current in many other infectious diseases with defined periodic *T* behavior (Earth seasonal (*ε*)) and we need to meet a periodic oscillation to predict *R*_0_ under *A*(*t*) criteria in time-varying environments with no heterogeneity forces, thus assuming a force of infection as 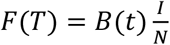 in order to be possible to establish a reasonable 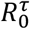 periodical stability for COVID-19 as observed by Bacaër [12] as defined in [12] as *p*(*t* + 1) = *A*(*t*) + *B*(*t*)*p*(*t*), being *p* the spectral matrix and *B*(*t*) the confounding environment (ecological variables such as biotic and abiotic) of compartments S, I and R of SIR model. At this point the seasonality of COVID-19 at S, I and R compartments is assumed to be dependent on deterministic outcomes for immunity, healthcare interventions and public policies under atmospheric triggering conditions (Earth seasons *ε*) as found, for example, in common flu. If considering this condition, the ODE could be easily observed in linear time series as pointed in Sietto [13] as 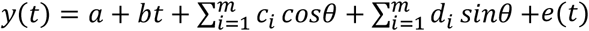, where the proposition of periodicity *θ* as linear in time as *B*(*t* + *T*) = *B*(*t*) would be possible and consistent in its fluctuations in terms of daily new infections with seasonal sinusoidal patterns as *θ*(*t*) = *θ*_0_[1 ± *ε* sin(2*πt*)] [14], and also stochastic over time factor considering seasonal fluctuations defined as Hidden Markovians chains as *P*(*Y*(*t*) = *y*(*t*) *Y*(*t* − 1) = *y*(*t* − 1), *Y*(*t* − 2) = *y*(*t* − 2), …, *Y*(1) = *y*(1)) [13] and its many derivations, found in many researches, as examples [15-17], of the same event worldwide that would lead to the seasonal Fourier transform fluctuations of COVID-19 outbreaks and over defined time behavior. If each epidemics is universally assumed as equal towards *ε* worldwide, then Fourier analysis would be possible to be performed considering time periodic fluctuations as noted in Mari *et al* [14] and therefore, the use of Markovian chains to obtain the phase shifts of regularities would be a true approach to predict how SARS-COV-2 spreading patterns are formed. The main problem we face here is when the stochastic process *Y*(*t*) assumes a lack of synchrony due to random delays [8,18,19] worldwide, hence generating a stochastic form with unknown seasonality of infection as defined in [18] as 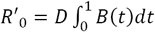, and therefore, not assuming a seasonality for *ε* and the outbreak of local epidemics. At this point we have several discrepant (heterogeneous) time series of the exponential behavior of daily new cases infection in countries that were in winter season (figure 1) and also comparing countries that are entering winter at south hemisphere and entering summer at north hemisphere (figure 1). There is no strong difference between Earth seasonality influencing those localities in its virus spreading patterns.

**Figure 1.**
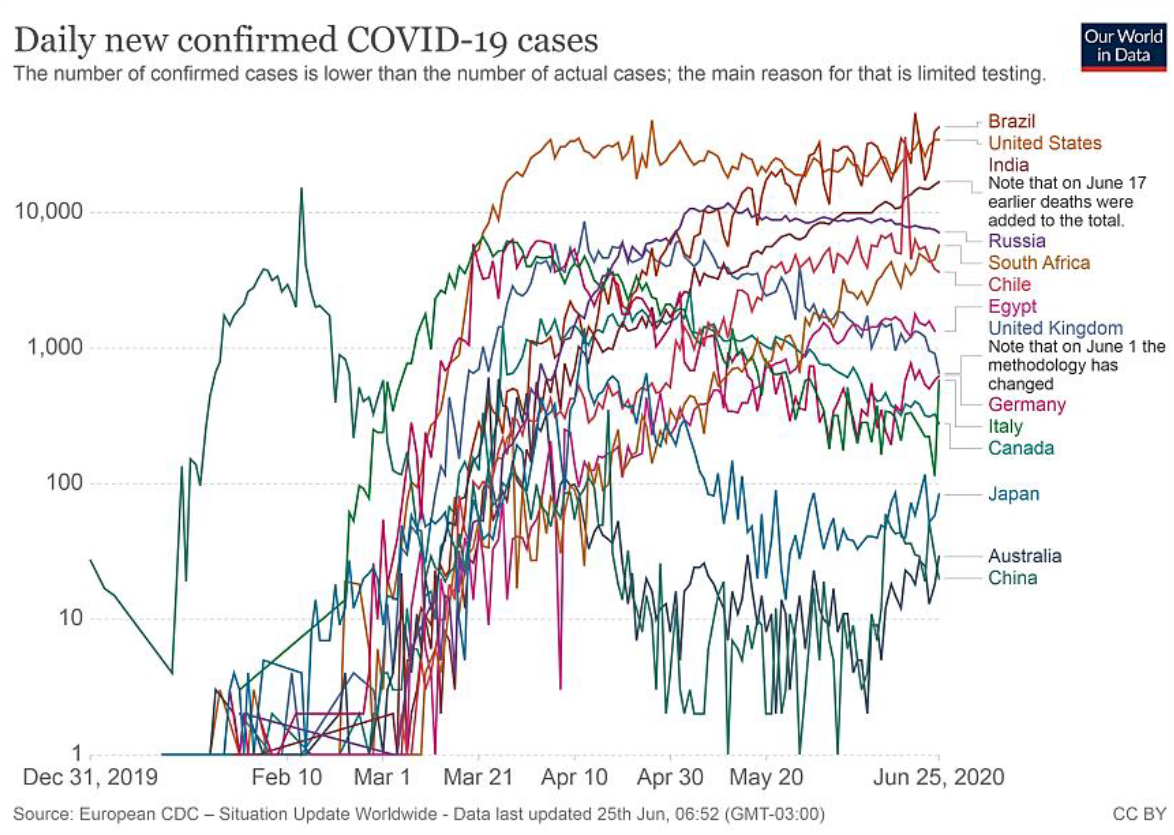
Selected countries from December 31, 2019 to June 25, 2020 from continents Asia, Europe, South and North America, Africa and Australia were displayed interpolating Earth seasons and number of cumulative daily new infections. Source: Our World in Data.

This lack of pattern formation as found in common flu [20] (figure 2 and 3) creates an undefined *T* over defined *A*(*t*) as well as mean *μ* (figure 2) over periodicity *θ* criteria (figure 3) as a pre assumption of analysis in the view of Fourier transform and therefore confirming an unexpected seasonality forcing behavior *ε*′ in which each sample (countries, regions, places,…) presents a different SARS-COV-2 spreading pattern not only concerning the Earth seasonality, but other components of *ε*′ as presented in the introduction section.

**Figure 2.**
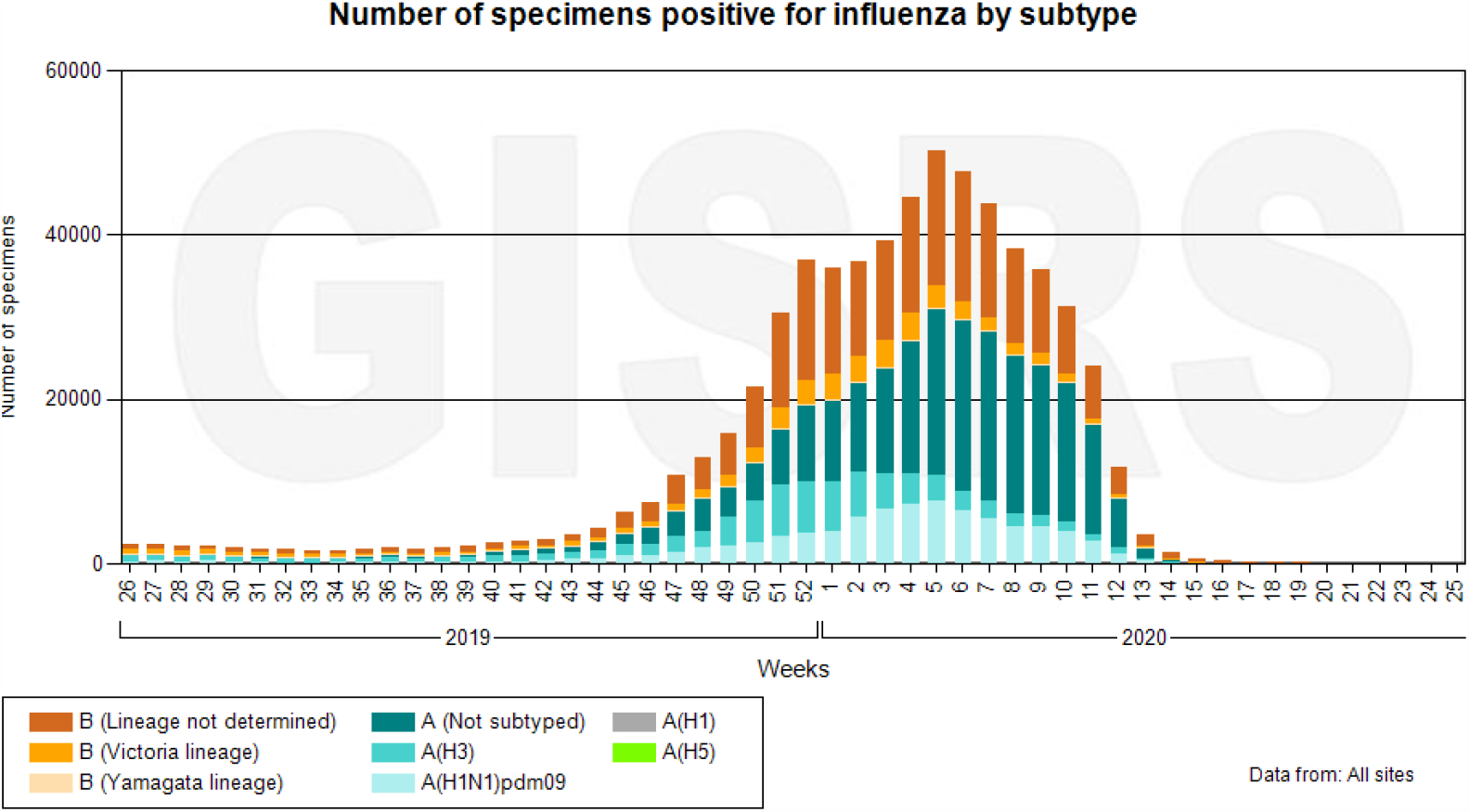
Seasonality of Influenza common species by 2019 and 2020 at North Hemisphere. Source: WHO.

**Figure 3.**
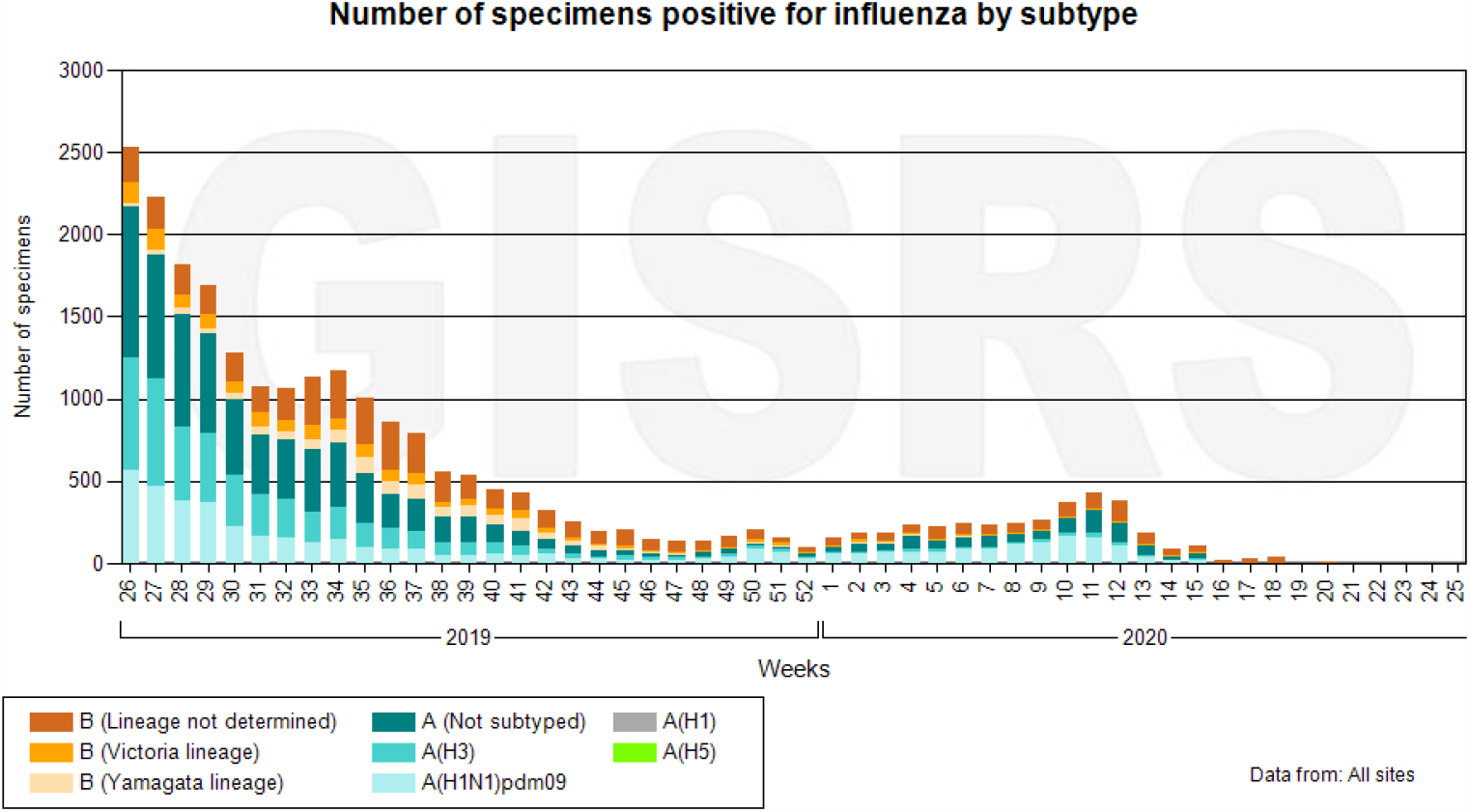
Seasonality of Influenza common species by 2019 and 2020 at South Hemisphere. Source: WHO.

### 2.2 Seasonality forcing behavior beyond Earth seasons and sinusoidal approaches

What we are observing clearly or maybe apparently at many results [2], is an asymptotic unstable behavior of SARS-COV-2 towards atmospheric conditions dictated by temperature, humidity, UV (ultraviolet) and wind speed as those results don’t present consistent indication of how atmospheric events have strong influence in the SARS-COV-2 spreading patterns of transmission as seasonal environmental drivers.

One might understand that north hemisphere had its first wave because of atmospheric conditions such as Earth season periods and for that in its turn, the south hemisphere as entering in winter will present exactly the same epidemics behavior presented at the first wave impact as observed in north regions. One important observation over it refers to the high amount of infection in north hemisphere in some countries under the summer season in contrast to the same amount of infection in south hemisphere under winter season that is occurring nowadays.

Both planet regions presented similar daily new infections at both seasons, therefore, not differing in the transmission spreading patterns of infection. Following this path, no periodic criterion was met for basic reproductive number *R*_0_ stability for an endemic equilibrium. We are facing daily new cases worldwide (figure 4) and the reason for the north hemisphere for European and Asian countries reduces its spreading patterns in the end of winter season is rather a coincidence over time that was caused mainly due to policies and ALE over population and individual behavior [3-8].

**Figure 4:**
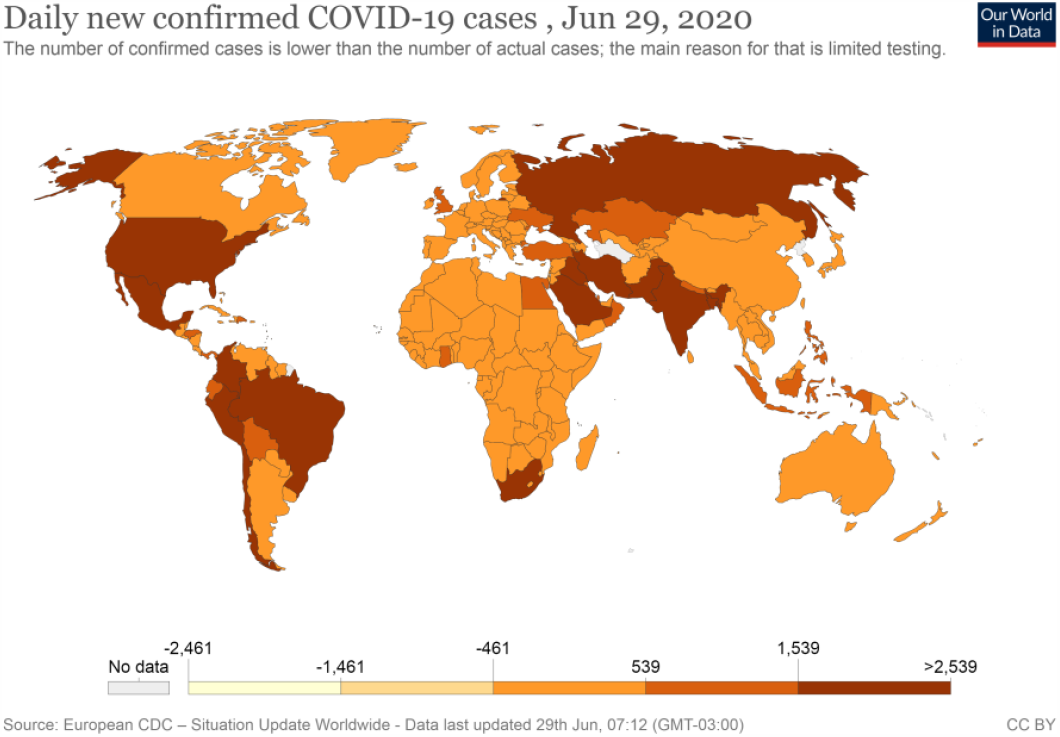
This graph mainly is focused in the comparison of north and south hemisphere countries such as Europe that decreased epidemics due to policies intervention and south and North America countries that were hit by first wave and policies were applied in different perspectives if compared to Europe. Source: Our World in Data.

If no seasonality of atmospheric conditions was found, we can observe still a seasonality forcing behavior, which was very well shaped by contact rates frameworks [4] based on policies and ALE actions [6] as represented in Figure 5. Bell shaped curve scheme. This overall scenario of pandemics could be very well observed in late March and starting April when at that time China and South Korea were the countries with the most lower rates of exponential growth of infection while Europe was in its growing pattern fully active and also in later June, many other researches pointed to the importance and role of policies and ALE towards pandemic control rather than atmospheric patterns of infection spreading worldwide [2-9].

**Figure 5:**
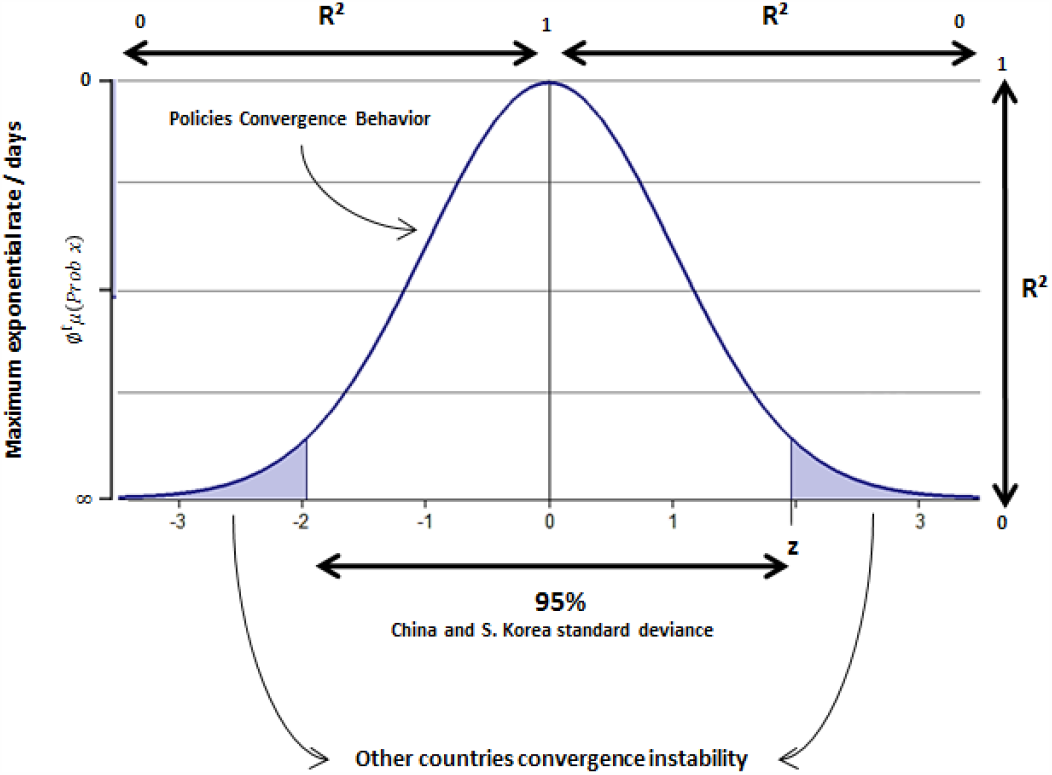
China and South Korea policies intervention effect over COVID-19 spreading pattern in late March and April, followed by other countries patterns. This was proved to be true [2-8] over time of pandemic outbreak regarding policies and SARS-COV-2 spreading patterns data.

But it does not mean that environmental variables such as atmosphere properties or Earth seasonality present no causation on the event (this will not be demonstrated and it is only theoretically assumed for the long-term expression of the pandemics, of which we don’t have still a visible glance of it). It means exactly that policies and ALE influence the phenomenon in different degree of seasonal forcing behavior than was expected to be addressed to the environmental factors, since we already have these outcomes available in worldwide data.

And also, not mentioned yet, the urban spaces found in every city, present specific potential to influence the local epidemics for the S and R compartments of SIR models, since it affects the capability of each country/city/locality to deal with the outcomes of susceptibility, immunity and public health control measures, therefore, making COVID-19 predictive models to assume data that are not perfectly real. And for each predictive model that fails to address urban spaces heterogeneity, policies and ALE interventions subjectivity and environmental non-homology of data, uncertainty degree grows making SARS-COV-2 emerge under unknown patterns of contagion as observed in Billings et al [19] and with a similar example of measles in Grenfell et al [21].

### 2.2 Seasonality forcing behavior

The unexpected seasonality under heterogeneity forcing behavior *ε*′ might confer to the exponential behavior of infection spreading patterns among countries an unpredictable sinusoidal expression like *β*(*t*) = *β*_0_(1 + *ε*Ø(*t*)) as pointed by Buonuomo et al [22] of Fourier transforms considering finite time lengths of analysis (seasons) equally distributed over time period *T* within samples (countries). This can be better understood because of the data series of cumulative daily new cases present high-amplitude noise and this is often related to the lower spectral density and lower frequency in which makes the analysis imprecise as a sinusoidal behavior in the basis form of Earth seasonality as 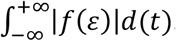. In this sense, the sinusoidal behavior does not exist in terms of how countries might present default oscillations within seasonal periods of Earth as represented schematically in figure 6.

**Figure 6.**
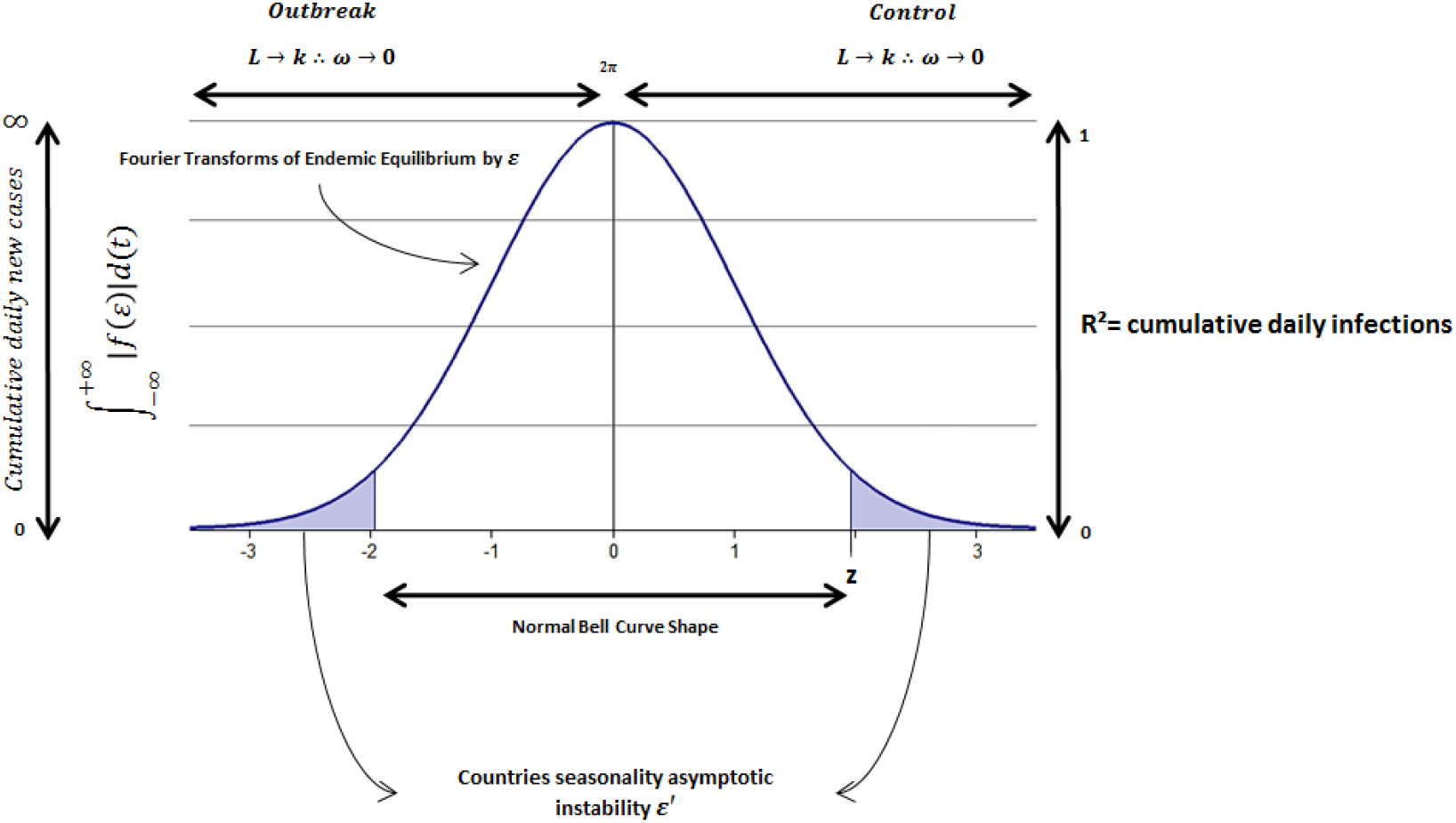
General framework of covid-19 seasonality under the view of Fourier transforms limitations.

And following this path, this leads to the observation that each sample can be understood as the lack of forming patterns towards confident interval and standard deviation under default time periods *T* from December 31, 2019 to June 25, 2020, resulting into a stochastic maximum exponential form of cumulative daily new infections as *Y*(*t*) change over time as showed in figure 7 samples.

**Figure 7.**
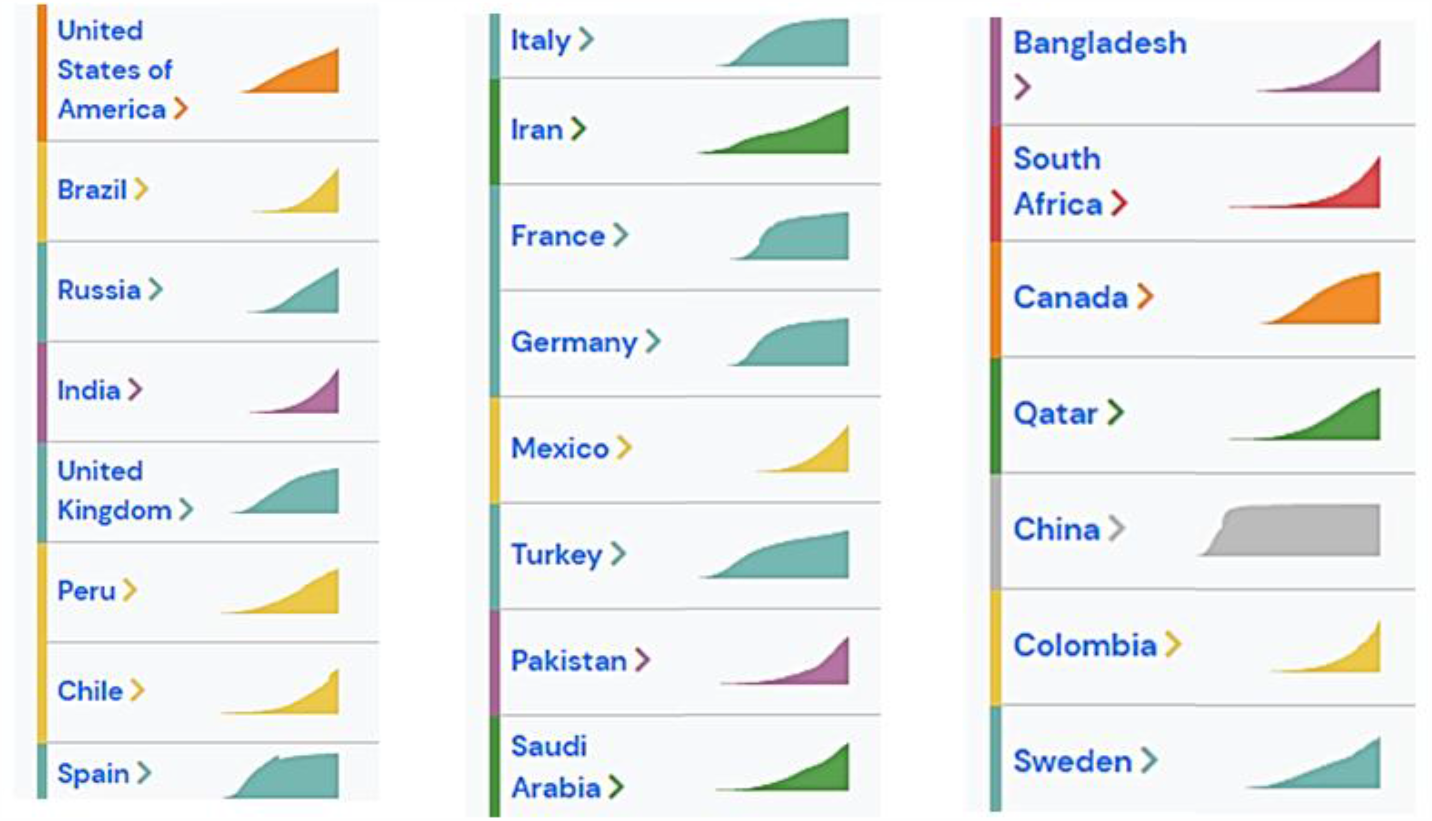
Some countries spreading patterns since outbreaks until June 25, 2020. Source: Outbreak.info.

However, despite of this scheme pointing to the weaker Earth seasonality forcing behavior of SARS-COV-2 spreading patterns, they can still be influencing the overall pattern of transmission with a hidden pattern due to policies/ALE interventions, environmental driven seasonality and urban spaces.

This point can be addressed to the pattern formation of *ε*′ confounding forced seasonality for S and R compartments over time [1,23-29], environmental driven factors [30-32] and policies/ALE intervention [3-8]. It is possible to observe (figure 1, 3 and 7) that each country dimension might respond differently to the same initial conditions, influenced by these three components, thus generating multiple patterns formation over time *T* for SARS-COV-2 transmission and periodicity.

Concerning a theoretical desired worldwide normal distribution that most mathematical models implies for infection spreading patterns with shape behavior *k* = 1 or *k* > 1 (Weinbull parameterization) of exponential “irregular” distributions of SARS-CoV-2 infection within time intervals *t* with defined periodicity *T* (seasonality among countries) [33], the defined original form of *I* compartment is given as 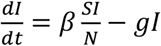. However, the high asymptotic instability behavior [23-29] of *I* lead us to redefine the equation basic fundaments and it can be understood as 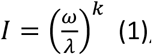 (1), where the infected *I* is influenced by unpredictable scale of infection *λ*(*N*) with inconsistent behavior of variables transition rate (*βSI*) defined as *ω*, and is not assumed for *gI* in the original form of R, that there are a normal distribution output for this virus spreading patterns. This new pattern formation of the epidemic behavior was well pointed by Duarte et al [34] when contact rate do not take in account weather conditions and time-varying aspects of epidemics. Therefore it was used an unpredictable shape *k* (close to reality shapes), mainly defining this shape caused *λ* and *ω* asymptotic instabilities generated by S and R compartments over time [1,23-29], environmental driven factors [30-32] and policies/ALE intervention [3-8]. This equation represents the presence of confounding and heterogeneous environmental variables *ω* with unknown predictive scale of *exp λ* or maximum likelihood estimator for *λ* due to nonlinear inputs for S and R (urban spaces), policies and environmental conditions influence, and therefore generating nonlinear outputs *k* (asymptotic instability) [35,36]. If we consider that most models are searching for a normality behavior among countries, hence, implying that the *k* distributions are non-complex and not segmented by its partitions, therefore resulting into a linearity for the virus infection *I* over *Y*(*t*) and *t*, then the overall equation as described by Dietz *β*(*t*) = *βm*(1 + *Acos*(*ωt*)) [35] would be not reachable for any given time period of analysis considering the seasonality forcing behavior of SARS-CoV-2.

The outputs with heteroscedasticity and non-homologous form for *k* and *λ* can be modified to reach stable points of analysis in as modeled by Dietz *β*(*t*) = *βm*(1 + *Acos*(*ωt*)) for each of the three seasonality forces influencing SARS-CoV-2 spreading patterns, that is regions where Fourier transforms and other methods of predictive analysis based on SIR models and derivations, policies interventions and the main role of environmental variables towards outbreaks and waves restarting periods can be found. These stable points of asymptotic convergence can be observed in the scheme of figure 8.

**Figure 8.**
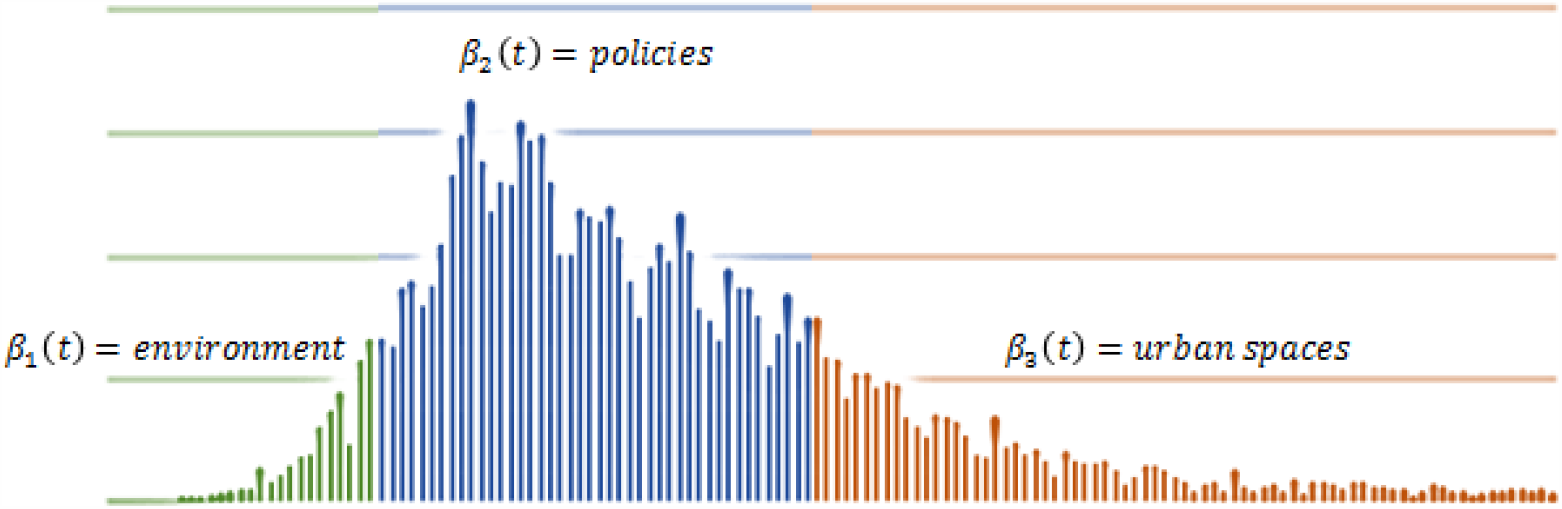
Concerning the apparent exclusion of Earth well defined seasonality, the external forcing behavior of SARS-COV-2 spreading patterns can be now filtered and stated as presenting three phases of expression, that is the S and R compartments constraints to modeling aspects, environmental driven and confounding variables and policies/ALE interventions conferring to the modeling aspects of prediction, undesirable uncertainty degree not only for outbreaks, second waves.

In order to remove heteroscedasticity and non-homologous form for *k* and *λ* from occurring, as far as the *κ* < 1 Weibull parameterization aspect [37] (Bell curve shape) of distribution be elected as the most reliable region of analysis (attractive orientation) for any given *T* periods within samples (countries cumulative daily new cases time series), it is necessary to modify the first equation (1) to 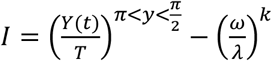 (2), hence with the new SIR model proposition as *I* = *I*′ − *S* + *R*, where *I* is asymptotic to *I*′ and S and R considered in its original form *θ*(*t*) = *θ*_0_[1 ± *ε*sin(2*πt*)] [14]. This is a mandatory redesign since many scientific breakthroughs are pointing to policies as the best approaches to reduce COVID-19 nowadays [3-8]. Starting with this redesign of equation we might find one of the first region of analysis and stability, which is policies intervention, found in the slope (peak) of daily cumulative cases over time.

Let’s address this persistence homology briefly for this research, where this desired mean function *Y*(*t*) of topological space 𝕩 → ℝ over *β*(*t*) = *βm*(1 + *Acos*(*ωt*)) indicated at (2) can be found as a persistence diagram existence [38] by mapping each adjacent pair to the point *f*(*Y*(*t*)), *f*(*t*) minimum and maximum observations, resulting in critical points of *Y*(*t*) function over time *t* not in adjacent form globally but regionally triangularly space as *d*(*D*(*Y*_*t*_), *D*(*t*) ≤ ∥*Y*_*t*_ − *t*∥_∞_ [39] with a given mean region, thus expressing random critical values defined by 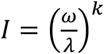 in the original form of observation of the event. But since we need to filter *f*(*Y*(*t*)) − *f*(*t*) unstable critical points (oscillatory instability of seasonality for S and R policies/ALE and environmental driven variables) to an attractive minimum behavior with normal distribution, then this region of analysis must be situated between 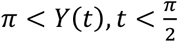 for every *A*(*t*) → *T* asymptote periods. Following this path, we going to have a roughly speaking the mean as the size of persistence diagram and triangulable diagonal (Δ) like 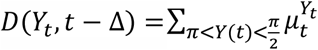 with multiplicity pairing regions (*t, Y*_*t*_) for each desired triangulation as 0 ≤ *t* < *Y*_*t*_ ≤ *n* + 1, resulting in the general equation for any assumed region as 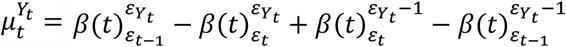 [39]. Note that each mean function 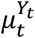 will be given by regions defined as *β*(*t*) = *βm*(1 + *Acos*(*ωt*)), being *β*(*t*) the covariance function of seasonality forcing behavior *μ*(*A*(*t*)), therefore without a global mean value for the event in terms of infection and time. Further derivations and formulations regarding this persistence diagram won’t be addressed for this research, but it is highly suggested that future researches keep these formulation defined for predictive and monitoring analysis of epidemic seasonality forcing behavior.

Also, it is necessary to understand that this new design of seasonality regions can be properly adapted to Fourier transform analysis under the amplitude of waves with the equation *e*^−*iωt*^ = cos(2*πt*) + *i sin*(2*πt*) where angular momentum was drawn in the limits of *β*(*t*) = *βm*(1 + *Acos*(*ωt*)), giving 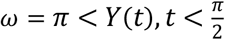 and generally defining it with sinusoidal reduced form as 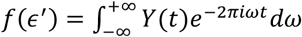 in order to reach a sinusoidal approach of time series data extraction and analysis over time periods and regions of analysis.

Also beyond the limitation of time periods for predictive analysis and monitoring as a Gaussian Process, this design also introduces one main point of analysis that is the lack of a mean and covariance function *μ*(*Y*(*t*)) over fluctuations as a global homomorphism and a decomposition form of wave signals similar to Fourier transforms where persistent homology can be found for *t* ∴ *κ* < 1 Weibull reliability to be situated in the oscillations pairing region of sin (*π*) = 1 and cos (*π*) = 0 for *T* desired coordinates of fluctuations in (*f*(*Y*_*t*_), *f*(*t*)) of stability with *t* + 1 continuous form as 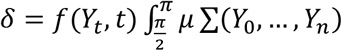*d_ε_* [9], thus assuming the shape and limit to *κ* < 1 as small partitions to the desired analysis or without a derivative form for the overall analysis within the period defined. For the discretized view of *Y*_*t*_, *t* as pointed in [9] results, it is possible to obtain a sample mean as 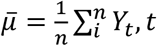. Further results of this approach can be visualized at [9] reference.

At this point, by rejecting the persistence diagram unstable critical points generated, a local minimum of the event as an average mean can be obtained by having *Y*(*t*) with the higher number of samples *Y* (daily infections) that finds a condition roughly described in the nonlinear oscillations within the exponential growth epidemic behavior of event as limited between local maximum growth defined by 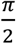 by its half curvature oscillations *π* as a local minimum being non periodic as 2*π* in a global homomorphism sense due to *κ* < 1. In this sense, the new sinusoidal approach offers new mean function as an angular momentum of 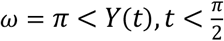. This scheme can be observed for policies/ALE intervention on SARS-CoV-2 spreading patterns [23] in figure 9.

**Figure 9.**
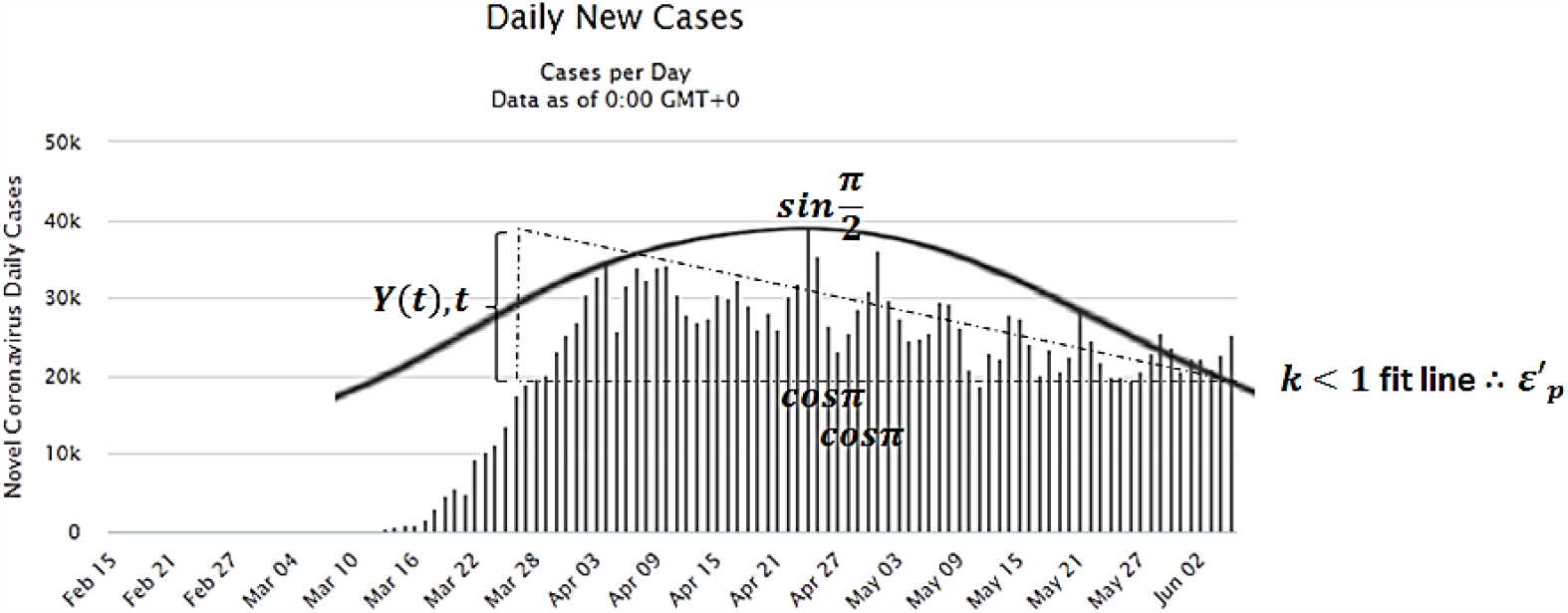
Policies/ALE stable region of analysis on SARS-CoV-2 spreading patterns. Image data source: Worldometer – Italy on 08 July 2020.

**Figure 9.**
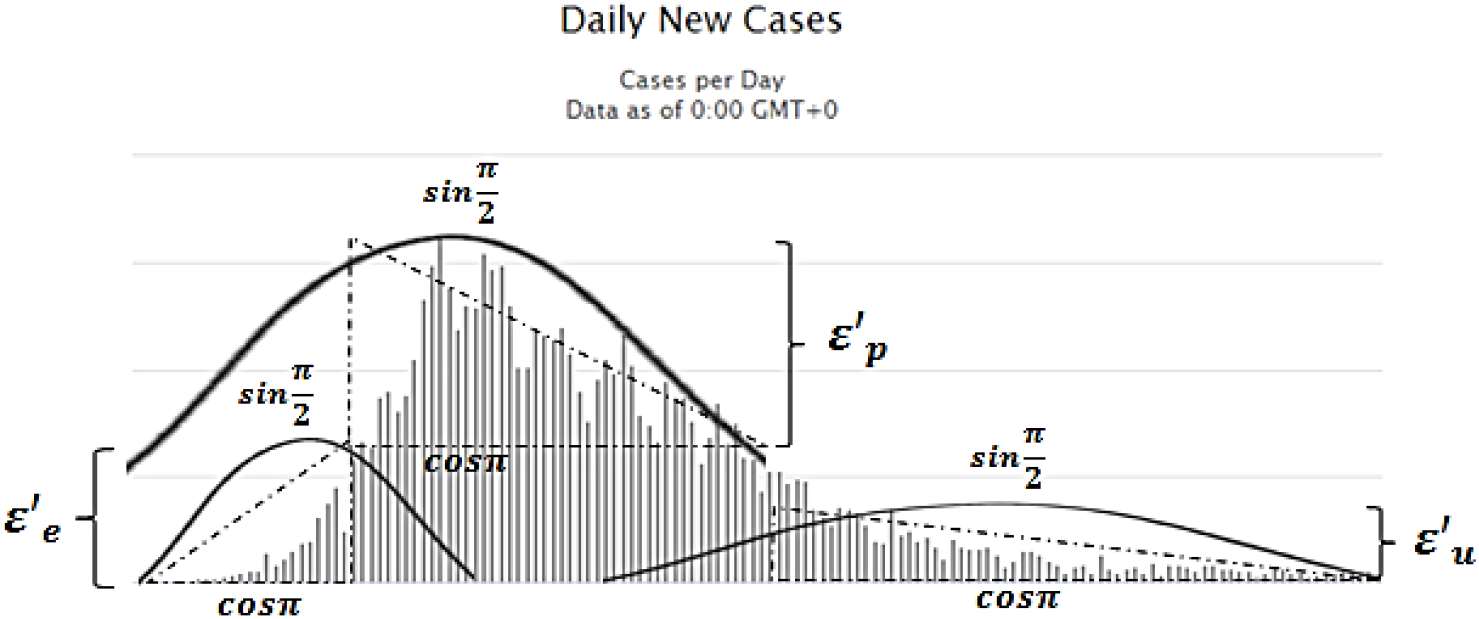
Considering the observation of figure 10, it is showed the asymptotic strong seasonality force of policies/ALE (*ε*′_*p*_) intervention and the narrow and instable region (outbreak and control) of analysis for environmental and urban driving factors of seasonality (*ε*′_*e*_, *ε*′_*u*_). Also regarding comportments S and R, the phase where control is reached by policies intervention remain as the most stable region of analysis for this SIR model equation compartments despite of instabilities cause by *ε*′_*e*_ and *ε*′_*u*_.

Therefore *Y*(*t*), *t* assumes the desired oscillations samples and region condition like 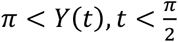 where persistent homology can be found for *t* ∴ *K* < 1 to be situated in the oscillations pairing region of sin (*π*) = 0 and cos (*π*) = *Y*(*t*) for *Y*(*t*), *t* desired coordinates (*f*(*Y*(*t*)), *f*(*t*)) of stability with *t* + 1 as 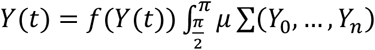 or vice-versa for 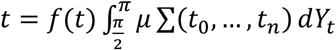, thus assuming the shape and limit to *κ* < 1. Then concerning time lengths of samples, it is designed as *t*(*δ* + 1) ≤ *f*(*Y*(*t*))*μ* Σ (*Y*_0_, …, *Y*_*n*_) *dt* starting from 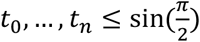 results in the desired data distribution with a conditional shape of Weibull parameterization *κ* < 1 for the analysis with a normal distribution, therefore rejecting any critical value beyond cos (*π*) = *ε*′_*p*_ and under sin (*π*) = *ε*′_*p*_, being *ε*′_*p*_ the seasonality forcing behavior of policies intervention over SARS-CoV-2 among countries data sets.

Now, considering that the S and R compartments of SIR model are needed to predictive analysis of infection spreading patterns, these compartments might work properly under the third region of time series data, that is the urban spaces *ε*′_*u*_ seasonality. To achieve these results with high reduction of uncertainty, it is necessary to conceive S and R as in its most stable region of analysis, that should be influenced in a posterior scenario where *ε*′_*p*_ (policies/ALE) and *ε*′_*e*_ (environmental seasonality) already took effect. This is mandatory since as far as policies are assumed in models or estimated with unreal quantitative parameters they promote uncertainty growth, and also they face limitations to track real patterns within an urban space features for S and R as a causation relation. For urban spaces seasonality forcing behavior, it is considered that inside and outside spaces promotes limitations to policies/ALE due to limiting action that it can face within these urban spaces (not all policies/ALE can survive in some urban spaces as it was designed to be). And also environmental seasonality can be present at this phase influencing with urban spaces the limitation of policies/ALE actions, therefore, *ε*′_*e*_ might find a spot to grow within inside and outside urban spaces beyond *ε*′_*p*_ normalization (more explanation of this causation effect will be given in results section).

Considering unexpected seasonal forcing *ε*′_*p*_ roughly defined as 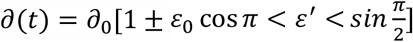 [9] in a complex network model, where non periodic oscillation (sinusoidal) are to be found in discrete form with 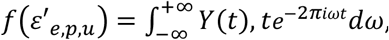, we might assume a rupture of the sin(2*πt*), leaving the region the pre assumed linearity *θ* (*t*) = *θ*_0_[1 ± *ε* sin(2*πt*)] for S and R in the overall metrics of time series data *T* within one sample or among countries and understand each iteration of the event as unconnected to the previous and future data if considering multiple time series comparisons (among countries) or even in the same time series if considering long-term analysis. In true, since the *I*′ is asymptote to *ε*′_*p*_, then *ε*′_*u*_ is limited by *ε*′_*p*_ on *I*′, but not necessarily fully stable in terms of *ε*′_*p*_ present total control over environmental seasonality due to urban spaces features. This statement is understood as far as policies/ALE interventions are the strongest attractive point of the phenomenon and therefore, compartment models find its limitation over how policies are implemented and how urban spaces can be convergent to policies/ALE interventions. Is is possible to check that most of these SIR models are constructed based on these *ε*′_*p*_ seasonality behaviors [40]. From this phase on, urban spaces and policies/ALE interventions might present high influence on the outcomes due to unpredictability of S and R patterns to design appropriate contact rate and that is still a limitation for the SIR model methods nowadays [23-29], however, it is still the most desirable region of analysis for data extraction.

Concerning urban spaces, due to huge diversity of public health infrastructure buildings design, outdoor and indoor building designs and natural physical features such as rivers, lakes, snow,…, it influences the environmental driven pattern on the region and not only policies. Therefore, it is reasonable to understand that any assumption on S and R during epidemic phase in its full curvature is much more closed to uncertainty measures than ever.

In this sense, countries might diverge seriously in the urban space and therefore, and S and R compartments finds limitations to calculate it during peak curvature and also it is contradictory since policies are not even fully developed or had the time to take effect while these models uses policies as the basis of modeling patterns. For this reason, the most reliable region of analysis for these compartments and where most of the models are situated/functioning actually, remains at the control phase of epidemics, that is when exponential behavior reaches asymptotic stability towards SARS-CoV-2 reduction, and of course, caused by policies interventions mostly. However, fluctuations still may occur worldwide due to the type of policy/ALE features and urban spaces features. In sense, no perfect prediction can be achieved still, but it is the most stable region for predictive analysis.

Now let’s assume that uncontrolled environmental driven factors *ε*′_*e*_ are the main cause of outbreaks and posterior waves of infection in a coupling relation with urban spaces and policies/ALE limitations, with unknown spreading pattern. This assumption can be confirmed since at this phase for the initial outbreak among countries, no policies intervention was existent or ALE features vary a lot among countries and also, as far as countries relax their policies [4,6,8], policies present urban spaces limitations and urban spaces promotes environmental seasonality, new waves of infection occur. In this sense, the environmental seasonality drivers as the main cause of aperiodic and unstable behavior for SARS-COV-2 spreading patterns worldwide.

Extracting the patterns of transmission of SARS-COV-2 at this point is challenging in terms of identify how outbreaks and positive control of epidemics occurs. It is possible to address to the outbreaks the main cause of environmental drivers of seasonality for SARS-COV-2 when we understand that no policies/distinct ALE are influencing the phenomenon at this phase. And also at this point of analysis, second or other waves of infection have its main focus for researchers since that, as outbreaks, waves can be very closed related to the environmental variables and urban spaces rather than any other form of seasonality forcing behavior.

The uncertainty growth of epidemics patterns worldwide within the outbreaks need to be understood excluding the policies intervention region of analysis, since this region present a stronger seasonality forcing behavior for SARS-COV-2 and therefore at this point, the environmental drivers and urban spaces will be hidden on its potential to influence the disease dynamics. Following this statement, the most reliable region to investigate environmental seasonality remains at the outbreak and control phases while urban space seasonality remain at control phase. This can be very useful for policies and ALE approaches since the fluctuations/instability present at this region is caused mainly by these two forcing behaviors and therefore, new strategies and measurements need to be adopted in order to keep economy and prevention with similar power.

It was observed lately that China presented a second wave of infection, as far as, it reduced some types of policies intervention, and therefore, it presented the urban spaces and environmental driving factor for SARS-COV-2 spreading patterns, being it the remaining infected citizens, environmental active virus, urban spaces outside scope of adopted policies or even the atmospheric influence for the disease transmission in any of these variables such as humidity, temperature, aerosols, wind, UV, etc.

If we could be capable of analyzing the outbreaks for first or second waves, we could be able to understand how SARS-COV-2 is influenced by urban and environmental seasonality comparing each country or region/locality with specific patterns for the environmental and urban variables. In this point, this research addresses new reformulations of urban and environmental variables influence and COVID-19 data sets under the view of cause and effect in the specific outbreaks region of analysis as showed in figure 10.

**Figure 10.**
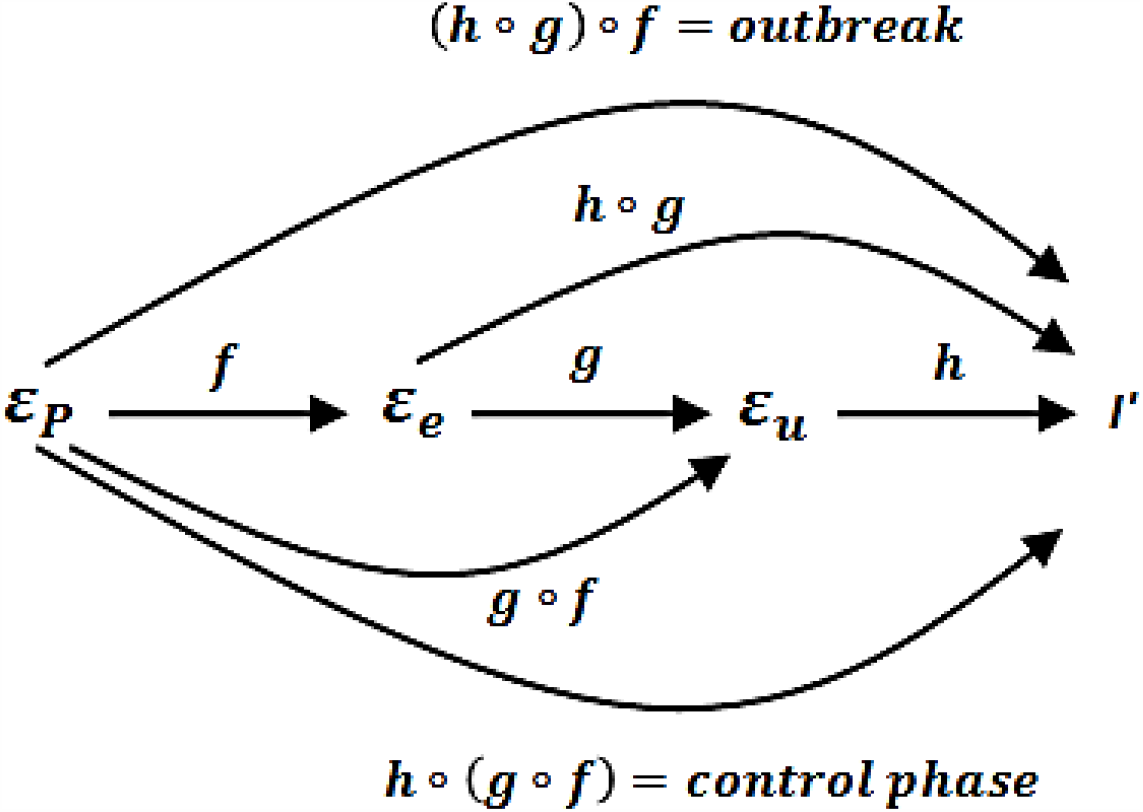
Schematically represented, the seasonality forcing behavior might assume the following behaviors: during the local epidemics, environment and urban spaces can be defined by policies/ALE until a limit; compartmental models are influenced by policies/ALE, environment and urban spaces; at control phase, policies/ALE finds its limitation by environment and urban spaces and finally, at outbreak, environmental factors present outcomes caused by the existing policies/ALE and urban spaces.

## 3) Results

The overall scenario of transmission and spreading patterns can be visualized by the scheme of figure 10, where seasonality forcing behavior assumes the following topological spaces. Considering all the possible seasonality types, 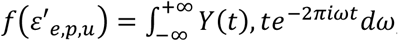, in continuous form of observation, and therefore, needing to be discretized within causal roots of analysis due to heterogeneity and confounding environment of analysis, therefore, each seasonality form can be understood as *fε*′_*p*_ = *g* ◦ *f*(*Y*_*t*_, *t*) = *g*(*f* (*ε*′_*p*_)), hence it can be also wrote as, *fε*′_*p*_ = *h* ◦ (*g* ◦ *f*(*Y*_*t*_, *t*)) = *h*(*g* (*f* (*ε*′_*p*_))) as a control phase of local epidemics. However, this phase might present high instability (fluctuations) worldwide due to heterogeneity and confounding behavior of *fε*′_*u*_ and *fε*′_*e*_. And since, SIR models need stable points for S and R, therefore *fε*′_*u*_ = *h* ◦ *g*(*Y*_*t*_, *t*) = *h*(*g*(*ε*′_*u*_)), resulting into a stable asymptotic convergence only if *fε*′_*p*_ = *h* ◦ (*g* ◦ *f*(*Y*_*t*_, *t*)) = *h*(*g* (*f* (*ε*′_*p*_))). And since the outbreak might find unknown patterns for *fε*′_*p*_, *fε*′_*e*_ and *fε*′_*u*_, then this region need to be carefully considered, and therefore environmental seasonality can be found as *fε*′_*e*_ = (*h* ◦ *g*(*Y*_*t*_, *t*)) ◦ *f* = *f*(*h* *g*(*ε*′_*e*_)) or it is also possible to assume *fε*′_*e*_ = (*ε* ◦ *g*(*Y*_*t*_, *t*), being *ε* the undefined patterns of environmental driven new infections for Earth seasonality or atmospheric factors.

Note that there are a great different between environment driven seasonality being caused by urban spaces influenced by policies/ALE limitations or otherwise caused by Earth or other natural (atmospheric) seasonality forcing behavior at outbreaks. This should be carefully considered when investigating Earth seasonality among countries. The compartmental models are mostly in the control phase region and they lose efficacy at outbreaks where no specific parameters are given and environmental seasonality is not yet discovered in its true patterns. Also, compartments models present high uncertainty during policies/ALE interventions phase and they work properly with empirical adopted policies (control phase) rather than pre assumed theoretical simulations. Another points is regarding control phase where instabilities occur as far as urban spaces creates a scenario where policies/ALE face limitations and therefore, environmental seasonality find suitable place to grow in its patterns.

Due to uncertainty growth over time and the lack of mean for defined intervals of *t* over *T* normal distribution shape for the whole data, Earth seasons *ε* loses its effect gradually as can be seen in figure 9 and 10 and the random delays observed for each country of analysis (sample) can be attributed by different patterns in which outbreak occurs since existing policies/ALE are found within world cultures, science and education. Earth seasonality should, for now, be addressed in terms of how it can influence transmission rather than seasonality patterns due to very limited overview of the event over time.

Also assuming the last researches on the field [3-8], policies and ALE are the most strong seasonality force influence the behavior of epidemics at second and third phase of time series data, while environmental factors are hidden in terms of transmission power at outbreaks and control phase giving unobserved and possible wrong results concerning this type of research. Another point of environmental convergence to SARS-COV-2 spreading patterns remains in the inside and outside of urban spaces where variables assume nonlinear forms of inputs and outputs and confounding outcomes with policies/ALE limitations, thus misleading the true behavior of infection spreading patterns under atmospheric parameters analysis (environmental seasonality). In this sense, it is very possible that environmental driven seasonality results found in many researches are in true, policies/ALE and urban spaces results. More detailed research need to be conducted. Inside and outside urban spaces are very important variables that can drive environmental research during all epidemics and concerning policies/ALE at the control phase and finally, possibly influencing with high uncertainty the compartment models for the susceptible and removed. For environmental seasonality evidences and complete description of atmospheric events influencing SARS-COV-2 transmission an artificial environment would be the best initial approach to reach that discover. This is because in the natural environment, the confounding variables involving transmission, presented as three phases in this research, can hide the true patterns of transmission considering UV, humidity, wind, temperature and other factors.

Therefore, having policies and ALE as the most convergent and stable interaction with the SARS-COV-2 spreading patterns, environmental and urban factors are presented as secondary influence which makes difficult for outside analysis to perform confident measures of its influencing power. The same happens for SIR models predictive analysis when it considers worldwide equal adopted policies or no environmental influence for the outbreak or control phases. Though secondary, it assumes a major importance at control phase, since it is the main cause of policies/ALE limitations to reduce or even end the transmission complex network.

One more seasonality forcing behavior that could be researched is about infodemics. This was not addressed in this research as a defined region within the time series data, since it is unpredictable in terms of empirical expression concerning individual and collective behavior phenomena. In other words, infodemics of subjective reasoning is something that occur momentaneously and independent of time or other static parameters. However it is predictable when it is adopted by policies as it occurred in some countries. Also it can be predictable if detected within a community by local authorities.

## 5) Conclusion

This research modeled patterns of SARS-COV-2 spreading by redesigning time series data extraction. This approach opened a new scenario where seasonality forcing behavior was introduced to understand SARS-COV-2 nonlinear dynamics due to heterogeneity and confounding scenario of epidemics where actual SIR models might find high degree of uncertainty. To overcome this limitation it was proposed the division of epidemics curvatures into regions where compartments of SIR models could be better understood and scientifically analyzed as well as pointing to the type of seasonality forcing behavior COVID-19 present worldwide.

Regarding curvature features, this research pointed to regions of analysis where seasonality forcing behavior of SARS-COV-2 finds it is most fitting causality for policies and ALE interventions, environmental driven factors and urban spaces. These regions were pointed as the most effective data for monitoring, control and predictive analysis.

Concerning the regions of analysis, not only Earth seasons, but atmospheric conditions (environmental driving seasonality) and urban spaces can present a transmission dynamics that is hidden in its pattern due to policies/ALE interventions worldwide, thus influencing predictive analysis of SIR models with uncertainty. However policies and ALE can be the strongest stable point of seasonality, it can find itself limited at control phase, depending on the region/country/locality given. This dynamics can be observed briefly, for the moment, in the random distributions of exponential behavior of countries where outbreaks of first and second waves are occurring as well as heteroscedasticity form of time series data worldwide.

Also, to overcome this hidden patterns scenario, it was found that seasonality forcing behavior can be tracked by new mathematical tools concerning data extraction among countries and within countries time period of infection. These tools can reveal new patterns formation regarding seasonality and therefore contributing to the future use of Fourier transforms in order to extract periodic phases of SARS-COV-2 transmission under specific set of parameters. Therefore, following these statements, it is very encouraged that researches in future adopt this angular momentum (regions of data extraction) of analysis for the environmental, policies/ALE and urban spaces patterns of transmission.

## Data Availability

Some of data used are available at Our World in Data and Outbreak website retrieved from: https://outbreak.info/data and https://ourworldindata.org.

## Author Contributions

Not applicable.

## Statement of Ethics

No humans or animals were involved in this study. Ethics approval was not required.

**No trial registration is applied for this study**.

## Acknowledgments

The author feels very grateful for the researchers that made this article possible through conversations and opinions about policies and ALE, by Henrique Lopes from the Association of Schools of Public Health in the European Region (ASPHER) and about environmental research by Manuel Hernández Rosales from Universidad Nacional Autónoma de México.

